# Enhancing Early Detection of Cognitive Decline in the Elderly: A Comparative Study Utilizing Large Language Models in Clinical Notes

**DOI:** 10.1101/2024.04.03.24305298

**Authors:** Xinsong Du, John Novoa-Laurentiev, Joseph M. Plasaek, Ya-Wen Chuang, Liqin Wang, Gad Marshall, Stephanie K. Mueller, Frank Chang, Surabhi Datta, Hunki Paek, Bin Lin, Qiang Wei, Xiaoyan Wang, Jingqi Wang, Hao Ding, Frank J. Manion, Jingcheng Du, David W. Bates, Li Zhou

## Abstract

**Background:** Large language models (LLMs) have shown promising performance in various healthcare domains, but their effectiveness in identifying specific clinical conditions in real medical records is less explored. This study evaluates LLMs for detecting signs of cognitive decline in real electronic health record (EHR) clinical notes, comparing their error profiles with traditional models. The insights gained will inform strategies for performance enhancement.

**Methods:** This study, conducted at Mass General Brigham in Boston, MA, analyzed clinical notes from the four years prior to a 2019 diagnosis of mild cognitive impairment in patients aged 50 and older. We used a randomly annotated sample of 4,949 note sections, filtered with keywords related to cognitive functions, for model development. For testing, a random annotated sample of 1,996 note sections without keyword filtering was utilized. We developed prompts for two LLMs, Llama 2 and GPT-4, on HIPAA-compliant cloud-computing platforms using multiple approaches (e.g., both hard and soft prompting and error analysis-based instructions) to select the optimal LLM-based method. Baseline models included a hierarchical attention-based neural network and XGBoost. Subsequently, we constructed an ensemble of the three models using a majority vote approach.

**Results:** GPT-4 demonstrated superior accuracy and efficiency compared to Llama 2, but did not outperform traditional models. The ensemble model outperformed the individual models, achieving a precision of 90.3%, a recall of 94.2%, and an F1-score of 92.2%. Notably, the ensemble model showed a significant improvement in precision, increasing from a range of 70%-79% to above 90%, compared to the best-performing single model. Error analysis revealed that 63 samples were incorrectly predicted by at least one model; however, only 2 cases (3.2%) were mutual errors across all models, indicating diverse error profiles among them.

**Conclusions:** LLMs and traditional machine learning models trained using local EHR data exhibited diverse error profiles. The ensemble of these models was found to be complementary, enhancing diagnostic performance. Future research should investigate integrating LLMs with smaller, localized models and incorporating medical data and domain knowledge to enhance performance on specific tasks.

## 1. Introduction

Large Language Models (LLMs), neural models with billions of parameters trained on extensive, diverse text corpora, have exhibited remarkable capabilities in clinical language understanding tasks.^1–5^ They offer distinct advantages over traditional rule-based and machine learning approaches, which are often trained from scratch on narrower clinical datasets.^6–8^ Previous studies showed that LLMs achieved impressive performance in a variety of clinical natural language processing (NLP) tasks such as question answering, named entity recognition, and information extraction^1,2^. However, the effectiveness of LLMs in identifying specific clinical conditions within real medical records remains less explored. Their lack of explicit training on specific medical records may affect their accuracy.^9^ This study aims to evaluate LLMs’ performance in detecting signs of cognitive decline within clinical notes. We use this as a use case to explore their effectiveness and compare their error profiles with those of traditional models trained on a domain-specific corpus. The insights gained from this study will inform strategies for further enhancement.

Alzheimer’s disease (AD) and related dementias (ADRD) affect millions of Americans,^10^ significantly reducing patient quality of life and imposing substantial emotional and financial burdens,^11^ with care costs projected to reach $1.1 trillion by 2050.^12^ Existing treatments offer only temporary relief,^13^ underscoring the urgent need for breakthroughs in AD/ADRD therapy.^14^ Timely detection of cognitive decline signs can facilitate early interventions and clinical trial involvement for AD/ADRD.^15–18^ Electronic health records (EHRs), particularly clinical notes, are critical resources for identifying early indicators of disease, yet traditional diagnostic tools and variability in screening practices complicate detection.^19–22^ NLP offers a promising solution by efficiently analyzing large datasets and identifying subtle signs of decline not easily captured in traditional diagnostics.^23^ Although studies have been conducted to identify cognitive decline using NLP,^7,24–26^ the effectiveness of LLMs specifically in identifying cognitive decline through EHRs remains under-explored.

This research utilizes LLMs within HIPAA-compliant computing environments for a pioneering exploration of EHR note analysis for cognitive decline detection. It evaluates the effectiveness and interpretability of LLMs compared to conventional machine learning methods and examines the synergy between LLMs and machine learning to enhance diagnostic accuracy. To the best of our knowledge, this initiative is the first of its kind to employ LLMs in this capacity, representing a significant innovation and contribution to the field.

## 2. Methods

### 2.1. Setting and Datasets

This study was conducted at Mass General Brigham (MGB), a large integrated healthcare system in Massachusetts, which has established secure, HIPAA-compliant cloud environments for deploying and evaluating LLMs with actual EHR data. Two LLMs were tested: the proprietary GPT-4^1^ via Microsoft Azure OpenAI Service API, and the open-source Llama 2 (13B)^2^ via an Amazon Elastic Compute Cloud (EC2) instance. Details on the cloud environments are provided in **Supplementary Material Section 1** and **Table S1**.

We utilized the same definition of cognitive decline and annotated datasets from a previous study.^19^ Cognitive decline encompasses various progressive stages, from subjective cognitive decline (SCD) to mild cognitive impairment (MCI) to dementia. It can be identified through mentions of signs, symptoms, diagnostic evaluations, cognitive assessments, or treatment details in clinical notes. Transient cases, such as memory loss due to medication, were labeled as negative for cognitive decline.

The annotated datasets comprised sections of clinical notes from the four years prior to the initial diagnosis of mild cognitive impairment (MCI, ICD-10-CM code G31.84) in 2019, for patients aged 50 years or older.^19^ Due to the low positive case rate across the sections, we used a list of expert-curated keywords (**Table 1**) to screen for sections likely indicating cognitive decline. **Table 2** shows that Dataset I, consisting of 4,949 keyword-filtered sections, was used to train two baseline models. For prompt development and LLM selection, 200 random samples from Dataset I (Dataset I-S) were used for performance assessment, while the remaining samples (Dataset I-A) were utilized for sample selection in prompt augmentation. Dataset II, which includes 1,996 random sections not subjected to keyword filtering, served for final testing.

**Table 1.**
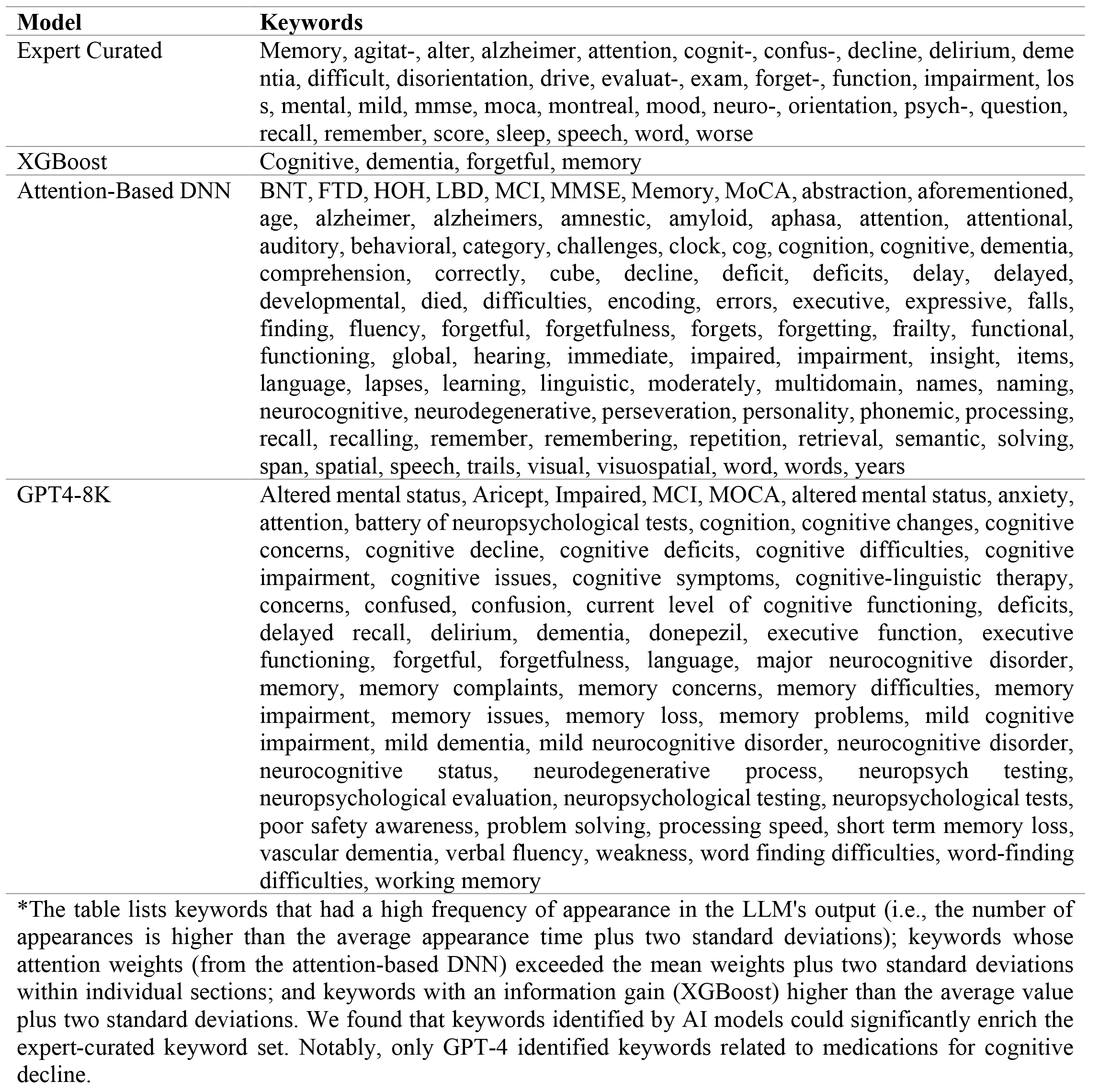
Keywords Contributing to the Identification of Positive Cognitive Decline Cases, Curated by Domain Experts and Extracted from AI Models.*

**Table 2.** Dataset Characteristics.

| **Dataset** | **Description** | **Mean (range),**<br>**characters** | **Positive**<br>**Rate** |
| --- | --- | --- | --- |
| Dataset I | 4,949 note sections filtered with cognitive decline-related keywords | 850 (26-9323) | 29.4% |
| Dataset I-A | A random subset of Dataset I, containing 4,749 samples | 848.8 (26-9323) | 29.6% |
| Dataset I-S | A random subset of Dataset I, comprising 200 samples that do not overlap with Dataset I-A | 870.7 (34-8353) | 23.5% |
| Dataset II | 1,996 random note sections without keyword filtering | 464 (26-14740) | 3.5% |

The study received approval from the MGB Institutional Review Board, with a waiver of informed consent for study participants due to the secondary use of EHR data.

### 2.2. LLMs and Prompting Methods

**Figure 1** (areas A and B) illustrates the two-step prompt engineering process: LLM selection and prompt improvement. Following previous studies, we divided the prompt into sections.^27^ Supplementary **Figure S1** shows the prompt structure, which includes a required task description and optional sections for prompt augmentation, error analysis-based instructions, and additional task guidance. We were cautious about the potential impact of longer prompts, which might overwhelm the model, negatively affecting performance, response speed, and cost efficiency.^28–30^. Therefore, as an initial step, we evaluated the performance of the two LLMs using manual template engineering and a smaller sample size. This approach enabled us to select the superior model and its corresponding prompt template for further analysis.^31^ The selection criterion was 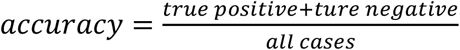, based on Dataset I-S. Using this metric and guided by the accuracy from Dataset I-S, we explored whether common prompt augmentation methods (both hard and soft prompting)^31^ and error analysis-based instructions^32^ could improve model performance. To ensure control over randomness and creativity, we adjusted the LLM’s temperature hyperparameter to 0, providing a deterministic solution.^32^

**Figure 1.**
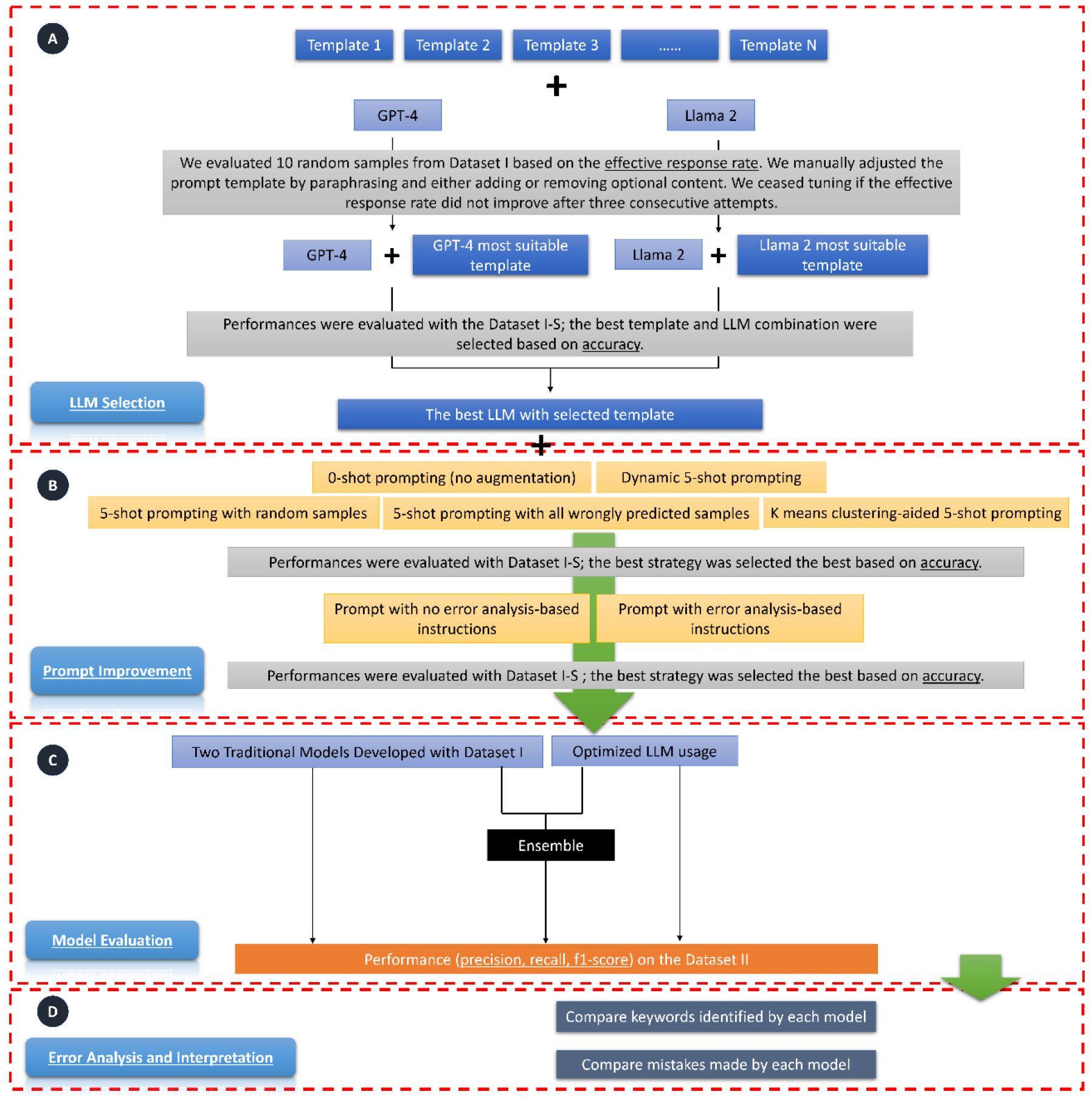
Study Design Overview. The workflow consists of five parts: A) LLM Selection: We fed prompts, which contain task descriptions and may also include additional task guidance as illustrated in Supplementary **Figure S1**, to GPT-4 and Llama 2 separately. We used 10 random samples from Dataset I to select the most suitable template for each LLM. During this process, if the effective response rate (i.e., the rate at which the response answered the questions in the prompt) was not 100%, we manually adjusted the template for each model. If the effective response rate did not improve after three consecutive attempts, we ceased tuning and used the template that led to the highest effective response rate. We then selected the best LLM based on their accuracy on Dataset I-S. B) Prompt Improvement: This step includes two sub-steps: prompt augmentation and adding error analysis-based instructions. During prompt augmentation, we tested whether five-shot prompting could improve accuracy. We then assessed whether incorporating instructions following an error analysis of the LLM’s output on Dataset I-S could enhance accuracy. C) Model Evaluation: We evaluated the selected LLM and two traditional machine learning models. We also tested the performance of an ensemble model, which took the majority vote of the three models as the predictive label. D) Interpretation and Error Analysis: For interpretation, we examined and compared keywords used by each model for prediction, in conjunction with those curated by domain experts. Lastly, we analyzed and compared errors made by each model.

#### 2.2.1. LLMs Comparison and Selection

We utilized an intuitive manual template engineering approach to fine-tune the task description and additional task guidance for each LLM.^31^ During the iterative refinement process, we focused on the following task descriptions for each LLM: 1) identifying evidence of cognitive decline in clinical notes; 2) displaying which keywords in the clinical notes informed its judgment on the assigned task; and 3) requiring LLM responses in JSON format to facilitate straightforward parsing. Furthermore, we explored the possibility of adding additional task guidance to assist the LLM in its reasoning and enhance performance. Specifically, we considered two approaches: 1) requesting the LLM to provide reasoning for its judgments, and 2) incorporating our definition of cognitive decline directly into the prompt.

##### Manual Template Engineering

We fed each prompt to GPT-4 and Llama 2 separately. The responses from these LLM were classified into three categories (Supplementary **Table S2**): 1) effective and parseable: the LLM’s response provides answers to both questions—whether cognitive decline was identified and which keywords were used for the decision—using a standard JSON format; 2) effective but not parseable: the LLM’s response answers both questions, but does not adhere to the standard JSON format; 3) not effective: the LLM’s response fails to answer either of the two questions. We assessed model effectiveness using 10 random samples from Dataset I. Our observations indicated that this sample size was sufficient for a meaningful comparison. If the effective response rate did not reach 100%, we manually adjusted the prompt template by paraphrasing or modifying optional content. This tuning process continued until no further improvement in the effective response rate was achieved after three consecutive attempts. Finally, we selected the prompt template that yielded the highest effective response rate for GPT-4 and Llama 2 separately.

##### Performance Comparison with Manually Crafted Templates

To select the optimal LLM, we compared the accuracy of GPT-4 and Llama 2 on Dataset I-S by providing the LLMs with manually crafted task descriptions and guidance.

#### 2.2.2. Prompt Improvement

##### Prompt Augmentation

We explored prompt augmentation to determine if including five examples (five-shot prompting) enhances performance. We adopted five-shot prompting due to the maximum token limitation of GPT-4. Since the selection of examples for few-shot prompting can significantly affect model performance^31,33^, we tested four different strategies, including both hard and soft prompting. To select the best strategy, we chose examples from Dataset I-A and evaluated model performance on Dataset I-S. The four example selection strategies were: 1) Hard Prompting - Random Selection: This strategy involves randomly selecting five samples. 2) Hard Prompting - Targeted Selection: We selected examples where the model had previously performed poorly, aiming to directly address its weaknesses. 3) Hard Prompting - K-Means Clustering-Aided Selection: This strategy involves selecting five samples from that are the centers of five clusters generated by k-means clustering. We utilized OpenAI’s embedding model, *text-embedding-ada-002*^33^, as features to ensure the examples are diverse and representative, which could be crucial for performance improvement. 4) Soft Prompting - Dynamic Selection: For each case in Dataset I-S, we automatically identified the top five most similar samples from Dataset I-A using OpenAI’s embedding model, *text-embedding-ada-002*,^33^ based on the k-nearest-neighbors algorithm. This process enabled us to provide the LLM with five samples that most closely resemble the current case, thereby guiding its decision-making.

##### Error Analysis-Based Instructions

We tested whether incorporating error analysis-based instruction into the prompt could improve performance.^32^ To achieve this, we first conducted an error analysis of the LLM on Dataset I-S. Subsequently, we added a paragraph describing common errors that the LLM made and instructed it to pay attention to those errors when generating its response.

##### Baseline Machine Learning Models

We compared the performance of the LLM with two baseline machine learning models developed from our previous study: XGBoost^34^ and a four-layer attention-based deep neural network (DNN),^7,35^ which incorporated elements of a convolutional neural network, a bidirectional long-short term memory (LSTM) network, and an attention model. These two models were the top performers compared to other traditional models in identifying cognitive decline in clinical notes.^19^

### 2.4. Ensemble Model

Finally, we investigated whether an ensemble model that combines predictions from both the LLM and traditional machine learning models could achieve better performance. The ensemble learning, which involves combining several different predictions from various models to formulate the final prediction, has proven to be an effective approach for enhancing performance.^36,37^ To create the ensemble model, we determined the label by taking the majority vote from the LLM, the attention-based DNN, and XGBoost. The high diversity of the models included may enable the ensemble to correct errors made by individual models.^38^

### 2.5. Model Evaluation

We evaluated and compared the selected LLM, traditional models and the ensemble model on Dataset II using standard metrics: 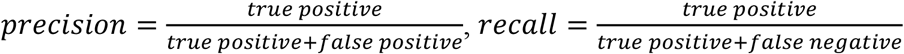, and 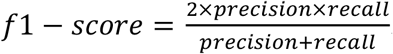. We used 0.5 as the cutoff point for calculating precision, recall, and F1 score for the baseline models.

### 2.6. Interpretation

Regarding interpretation, we listed keywords from the LLM’s output that appeared more frequently than the average appearance time plus two standard deviations. We also identified keywords whose deep learning attention weights exceeded the mean weights by more than two standard deviations within individual sections, and keywords with an XGBoost information gain higher than the average value plus two standard deviations. Additionally, we included expert-curated keywords developed in our previous study as a reference.^19^

### 2.7. Error Analysis

We conducted two levels of error analyses. The first analysis assessed the selected LLM using various prompting strategies, including zero-shot, the best few-shot method, and the prompt with error analysis-based instructions. The second analysis evaluated the best-performing LLM with its optimal prompt, alongside the attention-based DNN, and XGBoost. Errors made by each model were analyzed and discussed by two biomedical informaticians and a physician. We quantified unique and overlapping errors made by each model using a Venn diagram.

## 3. Results

Dataset characteristics are illustrated in **Table 2**. The average length of the Dataset I sections was 850 characters (range: 26-9393), and that of the Dataset II sections was 464 characters (range: 26-14740). Dataset I contained 29.4% positive cases and Dataset II contained 3.5% positive cases.

### 3.1. LLM Selection and Prompt Selection

The effective response rate varied for each LLM using five different prompt templates (**Figure 2**). For GPT-4, Template 1, which includes a task description section and additional task guidance section as shown in Supplementary **Figure S1**, achieved a 100% effective response rate. Llama achieved its highest effectiveness at 80% when using Template 2, which only includes the task description section. GPT-4 and Llama 2, with their most effective prompts, achieved accuracies of 86.5% and 52.0% respectively on Dataset I-S. We therefore chose GPT-4 for subsequent analysis.

**Figure 2.**
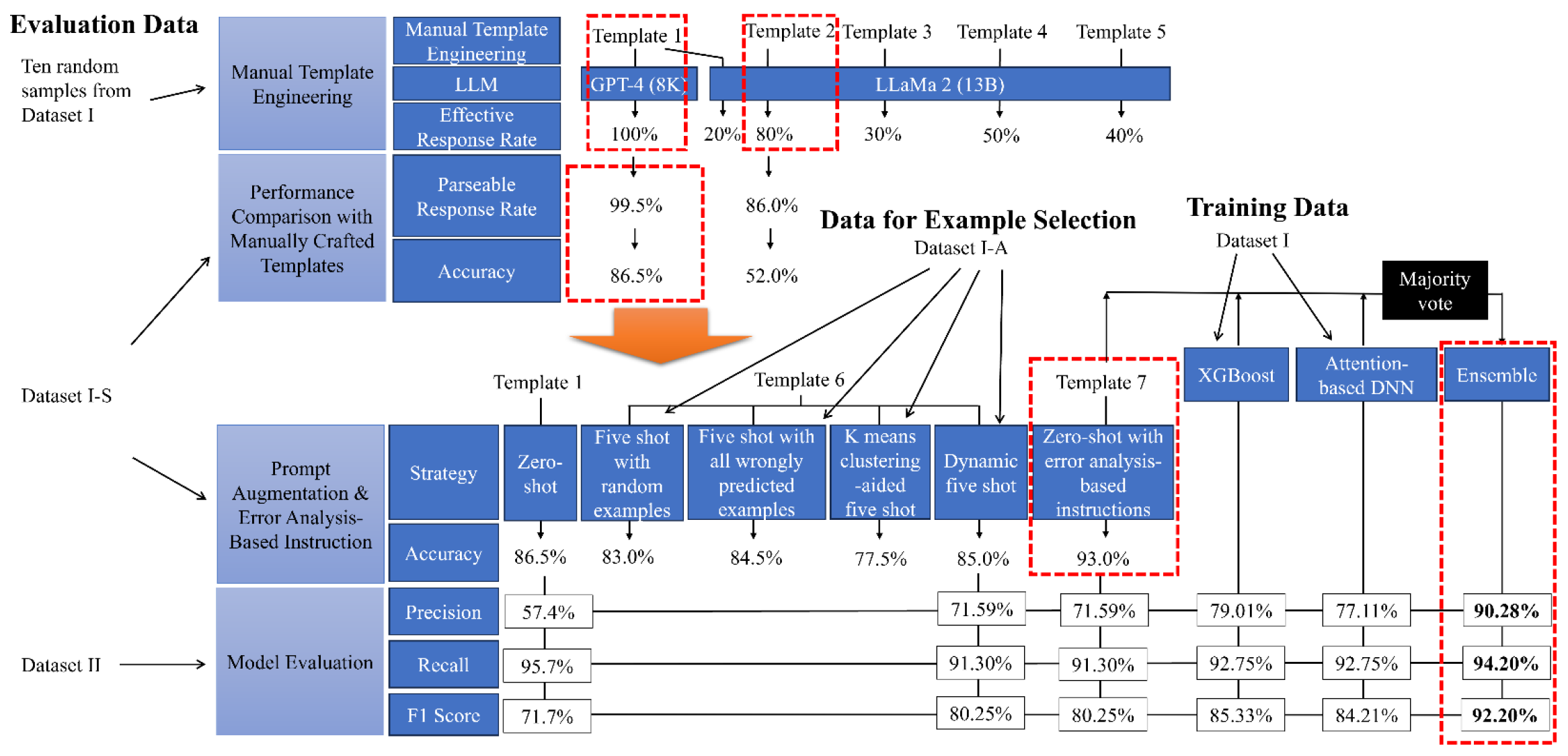
Evaluation Results Summary. During the prompt template selection, Template 1 was selected for the GPT-4 model due to a 100% effective response rate; Template 2 was selected for the Llama 2 model as the effective response rate (80%) did not improve after three tuning attempts. Subsequently, we compared the two combinations with 200 samples from Dataset I-S and found that GPT-4 and Template 1 combination achieved significantly better accuracy (86.0%). We also discovered that five-shot prompting did not lead to improved performance; however, adding error analysis-based instructions (i.e., GPT-4 and Template 7 combination) increased the accuracy to 93% on Dataset I-S. Consequently, we opted to use Template 7 as the prompt template and GPT-4 as the LLM. In tests, we evaluated the performance of the XGBoost, the attention-based DNN, and the LLM. We found that XGBoost performed better: precision – 79.01%, recall – 92.75%, and F1 score – 85.33%. Notably, after assembling the three models using a majority vote, the ensemble model demonstrated significantly improved performance: precision – 90.11% (an 11.1% improvement), recall – 94.20% (a 1.45% improvement), and F1 score – 92.20% (a 6.87% improvement).

Prompt improvement result on Dataset I-S shows that the best prompt augmentation approach (Template 6) was soft prompting – dynamic five-shot, which had an 85% accuracy. However, adding error analysis-based instructions (Template 7) surpassed this, reaching an accuracy of 93%. Therefore, we decided to adopt error analysis-based instructions as our prompting strategy for subsequent analyses.

### 3.2. Performance Evaluation

GPT-4 achieved a precision of 71.6%, a recall of 91.3%, and an F1 score of 80.3%. Optimized hyperparameters for attention-based DNN and XGBoost are illustrated in Supplementary **Table S8**. Attention-based DNN achieved a precision of 77.1%, recall of 92.8, and F1 score of 84.2%. XGBoost model achieved a precision of 79.0%, recall of 92.8%, and F1 score of 85.3%. Notably, the ensemble model significantly improved overall performance, achieving a precision of 90.3%, a recall of 94.2%, and an F1 score of 92.2%.

### 3.3. Interpretation

**Table 1** contains keywords identified through expert curation and exported by GPT-4, the attention-based DNN, and XGBoost. These keywords encompass a range of topics, including memory-related issues such as recall and forgetfulness, cognitive impairments, and dementia, with terms like ‘dementia’ and ‘Alzheimer’s.’ They also cover evaluation and assessment methods, referencing tools like the MoCA and MMSE. Compared to traditional AI models and expert-selected keywords, GPT-4 highlighted specific treatment options, notably ‘Aricept’ and ‘donepezil,’ (Supplementary **Table S9**) which are important in managing dementia and Alzheimer’s disease. Furthermore, GPT-4 explicitly identified specific diagnoses or conditions more than other models, with terms such as ‘mild neurocognitive disorder,’ ‘major neurocognitive disorder,’ and ‘vascular dementia.’ Additionally, GPT-4 exported keywords regarding the emotional and psychological effects of cognitive disorders, such as ‘anxiety,’ thus addressing aspects sometimes overlooked by other models.

### 3.4. Error Analysis

As illustrated in Supplementary **Figure S2**, when using different prompting strategies with GPT-4, some errors may be mitigated, while new ones could emerge that were not previously observed. Notably, adding error analysis-based instructions to the prompt yielded the best performance, with only 31 wrongly predicted cases in Dataset II. In contrast, the error profiles of GPT-4, attention-based DNN, and XGBoost exhibited much higher diversity (**Figure 3)**. We found that 63 cases were wrongly predicted by one or more models. GPT-4 accounted for 31 incorrect predictions, the attention-based DNN made 23 wrong predictions, and XGBoost was responsible for 22 incorrect predictions. However, only 2 (3.2%) cases were wrongly predicted by all models. Four errors were common between GPT-4 and the attention-based DNN, three were common between GPT-4 and XGBoost, and eight were shared between the attention-based DNN and XGBoost.

**Figure 3.**
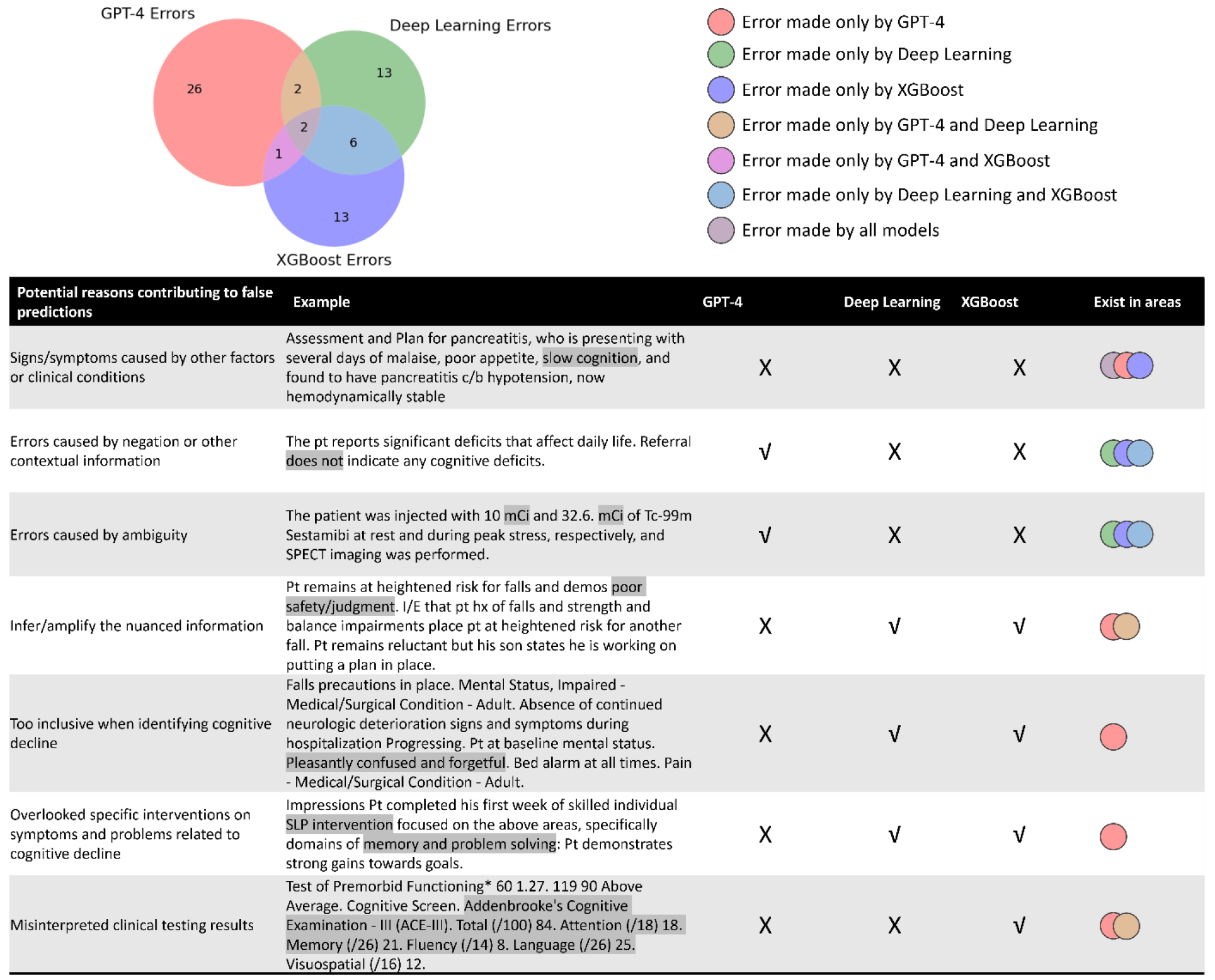
Venn Diagram Highlighting Unique and Overlapping Mistakes Made by Different Models. √: correct prediction; X: incorrect prediction. Some important findings include: 1) All models were susceptible to misinterpreting signs or symptoms as indicative of unrelated clinical conditions. 2) GPT-4 excelled in handling ambiguous terms and interpreting nuanced information, challenges that traditional AI frequently encounters. 3) Unlike traditional models, GPT-4 handles negations and contextual details more efficiently. 4) However, GPT-4 could sometimes overinterpret nuanced information or be overly conservative, failing to recognize whether a patient has cognitive decline despite strong evidence. 5) GPT-4 might also overlook certain medical domain knowledge, such as treatments or visits related to cognitive decline. 6) Both GPT-4 and attention-based DNNs occasionally misread clinical testing results, highlighting an opportunity for further improvement.

All models were susceptible to misinterpreting signs or symptoms as indicative of unrelated clinical conditions. GPT-4 excelled in handling ambiguous terms and interpreting nuanced information, a frequent challenge for traditional AI. Unlike traditional AI, GPT-4 was not confused by negations and contextual details. However, it could sometimes overinterpret nuanced information or be overly conservative, failing to recognize cognitive decline despite strong evidence. It might also overlook underlying causes of clinical events like treatments or visits related to cognitive decline. Both GPT-4 and attention-based DNNs occasionally misread clinical testing results.

## 4. Discussion

Recently, LLMs have demonstrated remarkable performance on various NLP tasks, yet their ability to analyze clinical notes from EHR data remains underexplored, partly due to data privacy concerns. In this study, we established HIPAA-compliant secure environments for LLMs and used cognitive decline identification as a use case to test LLMs’ capabilities in clinical note classification, thereby enhancing diagnostic tasks. Our contributions are threefold: 1) This study is the first to set up a secure cloud environment for GPT-4 and tested its ability to identify cognitive decline from clinical notes in EHR data; 2) We introduced a novel method for implementing NLP models for cognitive decline identification, achieving state-of-the-art performance with a significant lead over existing methods; 3) We discovered that although existing LLMs may not outperform traditional AI methods trained on a local medical dataset, their error profile differs distinctly, underscoring the significant potential of combining LLM with traditional AI models.

Our research demonstrated that prompt engineering using error analysis-based instructions significantly enhanced performance compared to zero-shot and prompt augmentation approaches, as it directly targeted the LLM’s weaknesses. Nevertheless, the LLM did not surpass traditional AI in identifying cognitive decline, primarily because it was not specifically trained for this task.^9,39^ While the LLM can generate a range of responses, it is prone to producing plausible but incorrect hallucinations. Nonetheless, it is valuable for its ability to operate without task-specific training, thereby complementing traditional AI, which requires specific training but often does not suffer from hallucinations.^40^ In terms of interpretation, the LLM identified keywords overlooked by experts and traditional AI models, such as medications related to cognitive decline. Error analysis revealed that the LLM demonstrated superior handling of ambiguous or contextually complex information due to its transformer architecture.^3,4^ However, LLMs misinterpreted or overlooked certain domain-specific medical tests and treatments. Future research should explore the integration of the LLM with smaller, localized models and knowledge bases to enhance performance on specific tasks.

Although our study has many strengths—for instance, it is the first to employ LLMs on unstructured EHR data for detecting cognitive decline—the results should be considered in light of several limitations. The LLMs used may not represent the most recent advancements (e.g., the recently released Llama 3 model) due to the rapid evolution of LLM technologies. While utilizing LLMs with a larger number of parameters (e.g., Llama 2-70 billion) may lead to better performance, this improvement comes with trade-offs, including higher computational demands and greater memory needs, posing challenges due to resource constraints. Additionally, our data are record-based and not patient-based (i.e., longitudinal), thus, the developed model may struggle to distinguish between reversible and progressive cognitive decline, and it remains unclear if patients recovered later based solely on a note from one time point. Therefore, developing an LLM-based early warning system for cognitive decline using longitudinal data would be a valuable direction for future research.

## 5. Conclusion

This study is among the first to utilize LLM within HIPAA-compliant cloud environments, leveraging real EHR notes for detecting cognitive decline. Our findings indicate that LLMs and traditional models exhibit diverse error profiles. The ensemble of LLMs and locally trained machine learning models on EHR data was found to be complementary, significantly enhancing performance and improving diagnostic accuracy. Future research could investigate methods for incorporating domain-specific medical knowledge and data to enhance the capabilities of LLMs in healthcare-related tasks.

## Data Availability

Due to the sensitive nature of the Electronic Health Records (EHR) data used in this study, direct access to the raw data cannot be provided. Where possible, aggregate data and summary statistics are available from the corresponding author upon reasonable request.

## Acknowledgments

This study was funded by NIH-NIA R44AG081006.

