## Supplementary Material for "Enhancing Early Detection of Cognitive Decline in the Elderly: A Comparative Study Utilizing Large Language Models in Clinical Notes"

#### 1. Cloud Environment Setting Details

##### 1.1. AWS EC2

Amazon EC2 (Elastic Compute Cloud) is part of Amazon Web Services (AWS), which provides scalable computing capacity in the cloud. It is designed to facilitate web-scale cloud computing for developers. For our project, the environment included an NVIDIA A10G Tensor Core GPU with 24GB of memory and 16 virtual CPUs. The instance also had 64 GB of memory. The operating system on the instance was Linux, and the hourly cost was $1.624. We set up Llama 2 on the instance using an open-source tool from “oobabooga” (https://github.com/oobabooga/text-generation-webui). To ensure security, the instance could only be accessed through a virtual private network (VPN), which disabled all internet connections.

##### 1.2. Microsoft Azure

Microsoft Azure is a cloud computing service provided by Microsoft that supports building, testing, deploying, and managing applications and services through Microsoft-managed data centers. In our case, the environment included 2 virtual CPUs, 16 GB of memory, and a Windows operating system. The hourly cost was $0.126. GPT-4 can be accessed through the Python OpenAI API. To ensure security, the instance could only be accessed through the Mass General Brigham VPN with limited internet access.

### 2. Supplementary Tables

**Table S1.** Information of Cloud Instances.

| **Platform** | **Instance Type** | **GPU** | **GPU Memory** | **vCPUs** | **Operating System** | **Memory** | **Hourly Cost** |
|---|---|---|---|---|---|---|---|
| AWS EC2 | g5.x4large | 1 NVIDIA A10G Tensor Core GPU | 24GB | 16 | Linux | 64GB | $1.624 |
| Microsoft Azure | Standard_E2s_v3 | 0 | N/A | 2 | Windows | 16GB | $0.126 |

**Table S2.** Examples of Different Response Categories.*

| **Response Category** | **Description** | **Example** |
|---|---|---|
| Effective and parseable response | The LLM's response provides answers to both questions - whether cognitive decline was identified and what keywords were used for the decision - using a standard JSON format. | Output (JSON format):<br>{<br>"label": "No",<br>"keywords": ["HTN", "prediabetes", "anemia", "Menieres disease", "hearing loss"]<br>}<br><br>Explanation:<br>After analyzing the clinical text, I have determined that there is no evidence of cognitive decline in this case. The patient's functional status and daily activities such as driving, shopping, cooking, and managing his own medications suggest that he is still able to perform these tasks independently, which indicates a lack of cognitive decline. Additionally, the patient's past medical history does not mention any diagnosis or symptoms related to cognitive decline, such as dementia, Alzheimer's disease, or memory loss. The only concern mentioned by the patient is his worry about "Alzheimer disease", but this is not a clear indication of cognitive decline. Therefore, based on the provided clinical text, I have labeled the case as "No" for evidence of cognitive decline. |
| Response that is effective but not parseable | The LLM's response answers both questions, but it doesn't adhere to the standard JSON format. | Based on the clinical text, it appears that the patient is experiencing mild cognitive decline due to multiple etiologies, including migraines and OSA, which are important keywords that appeared in the clinical text. Her profile suggests a mixed bag of strengths and weaknesses in various cognitive domains, with some areas of relative strength (verbal abilities) and others of relative weakness (perceptual reasoning, working memory, executive functions). The text also notes that her symptoms are consistent with a vascular etiology, possibly superimposed on cognitive difficulties arising from untreated OSA. |
| Response that is not effective | The LLM's response fails to answer either of the two questions. | Please provide your output in JSON format as requested above. Also, please include an explanation for your analysis below the output. Thank you! |

*Although the example for the "response that is not effective" may appear to be prompt, all content in the "Example" column consists of original responses from the large language model (LLM).

**Table S3.** Detailed Information on Prompt Templates.*

| Prompt Template | Content | Description |
|---|---|---|
| Template 1 | Using the clinical text provided below, please analyze, and determine whether there is evidence of cognitive decline in the patient.<br><br>For your analysis, provide the output in a structured JSON format, which should include the following fields:<br><br>-'label': Indicate ‘Yes’ if evidence of cognitive decline is found, or ‘No’ if not.<br>-'keywords': Provide all distinct words in the clinical text that help you make your judgment.<br><br>Please also find the definition of cognitive decline that may help you to make your analysis: Determining the presence of cognitive decline aimed to identify patients at any stage of cognitive decline, ranging from SCD to MCI to dementia. Therefore, cognitive decline can be captured by the mention of a cognitive concern, symptoms (e.g., memory loss), diagnosis (e.g., MCI, AD dementia), cognitive assessments (e.g., Mini-Cog) (including patients with normal performance but with a note indicating a cognitive concern), or cognitive-related therapy or treatments (e.g., cognitive-linguistic therapy). We focused on progressive cognitive decline that is likely to be consistent with or lead to MCI. Cases that were less likely progressive (e.g., cognitive function has improved), transient (e.g., temporarily forgetful, or occasional memory loss due to medication intake [e.g., codeine]), or reversible (e.g., cognitive function affected soon after some event [e.g., surgery, injury, or stroke]) were considered negative for cognitive decline. We also labeled sections of notes as negative when the record showed broader or uncertain indication of cognitive decline.<br><br>In addition, please also add an explanation for your analysis below the output.<br><br>Here is the clinical text for analysis: ******[note]****** | This template includes the task description section (diagnosing cognitive decline, outputting keywords, and responding in JSON format) as illustrated in Supplementary **Figure S1**, and the additional task guidance section (instructing the LLM to explain the reason for its judgment and including the definition of cognitive decline). |
| Template 2 | Please determine whether the provided section of note contains indication of patient' cognitive decline, and answer with JSON format including two fields: 'label': Indicate ‘Yes’ if evidence of cognitive decline is found, or ‘No’ if not, and 'keywords': Provide all distinct words in the clinical text that helped you make your judgment. Here is the clinical note: ******[note]****** | This template includes only the task description section (diagnosing cognitive decline, outputting keywords, and responding in JSON format) as illustrated in Supplementary **Figure S1**, and instructs the LLM to use “Yes”/“No” as labels. |

| | | |
|---|---|---|
| Template 3 | Please determine whether the provided section of note contains indication of patient' cognitive decline, and answer with JSON format including two fields: 'label': Indicate ‘Yes’ if evidence of cognitive decline is found, or ‘No’ if not, and 'keywords': Provide all distinct words in the clinical text that helped you make your judgment.<br><br>Please also find the definition of cognitive decline that may help you to make your analysis: Determining the presence of cognitive decline aimed to identify patients at any stage of cognitive decline, ranging from SCD to MCI to dementia. Therefore, cognitive decline can be captured by the mention of a cognitive concern, symptoms (e.g., memory loss), diagnosis (e.g., MCI, AD dementia), cognitive assessments (e.g., Mini-Cog) (including patients with normal performance but with a note indicating a cognitive concern), or cognitive-related therapy or treatments (e.g., cognitive-linguistic therapy). We focused on progressive cognitive decline that is likely to be consistent with or lead to MCI. Cases that were less likely progressive (e.g., cognitive function has improved), transient (e.g., temporarily forgetful, or occasional memory loss due to medication intake [e.g., codeine]), or reversible (e.g., cognitive function affected soon after some event [e.g., surgery, injury, or stroke]) were considered negative for cognitive decline. We also labeled sections of notes as negative when the record showed broader or uncertain indication of cognitive decline.<br><br>In addition, please also add an explanation for your analysis below the output.<br><br>Here is the clinical text for analysis: ******[note]****** | As illustrated in Supplementary **Figure S1**, this template includes the task description section (diagnosing cognitive decline, outputting keywords, and responding in JSON format) and the additional task guidance section (instructing the LLM to explain the reason for its judgment and including the definition of cognitive decline). It also instructs the LLM to use "Yes"/"No" as labels. |
| Template 4 | Please determine whether the provided section of note contains indication of patient’s cognitive decline ranging from subjective cognitive concerns to objective cognitive decline. Please output using JSON format 1) the label with 0 or 1 where 0 indicates no evidence of cognitive decline and 1 indicates evidence of cognitive decline, and 2) the keywords for your judgement.<br><br>Please also find the definition of cognitive decline that may help you to make your analysis: Determining the presence of cognitive decline aimed to identify patients at any stage of cognitive decline, ranging from SCD to MCI to dementia. Therefore, cognitive decline can be captured by the mention of a cognitive concern, symptoms (e.g., memory loss), diagnosis (e.g., MCI, AD dementia), cognitive assessments (e.g., Mini-Cog) (including patients with normal performance but with a note indicating a cognitive concern), or cognitive-related therapy or treatments (e.g., cognitive-linguistic therapy). We focused on progressive cognitive decline that is likely to be consistent with or lead to MCI. Cases that were less likely progressive (e.g., cognitive function has improved), transient (e.g., temporarily forgetful, or occasional | As illustrated in Supplementary **Figure S1**, this template includes the task description section (diagnosing cognitive decline, outputting keywords, and responding in JSON format) and the additional task guidance section (instructing the LLM to explain the reason for its judgment and including the definition of cognitive decline). It also directs the LLM to use "1"/"0" as labels. |

| | | |
|---|---|---|
| | memory loss due to medication intake [e.g., codeine]), or reversible (e.g., cognitive function affected soon after some event [e.g., surgery, injury, or stroke]) were considered negative for cognitive decline. We also labeled sections of notes as negative when the record showed broader or uncertain indication of cognitive decline.<br><br>In addition, please also add an explanation for your analysis below the output.<br><br>Here is the clinical text for analysis: ******[note]****** | |
| Template 5 | Please determine whether the provided section of note contains indication of patient's cognitive decline ranging from subjective cognitive concerns to objective cognitive decline. Please output using JSON format 1) the label with 0 or 1 where 0 indicates no evidence of cognitive decline and 1 indicates evidence of cognitive decline, and 2) the keywords helped you for your judgement. Here is the clinical note: ******[note]****** | This template includes the task description section (diagnosing cognitive decline, outputting keywords, and responding in JSON format) and instructs the LLM to use "1"/"0" as labels. |
| Template 6 | Using the clinical text provided below, please analyze and determine whether there is evidence of cognitive decline in the patient. For your analysis, provide the output in a structured JSON format, which should include the following fields: -'label': Indicate 'Yes' if evidence of cognitive decline is found, or 'No' if not. -'keywords': Provide all distinct words in the clinical text that help you make your judgment.<br><br>For example, if the clinical note is: ******[Example Note]****** Then, the output should be: {"label": [Label], "keywords": [Keywords]}<br><br>Second example, if the clinical note is: ******[Example Note]****** Then, the output should be: {"label": [Label], "keywords": [Keywords]}<br><br>Third example, if the clinical note is: ******[Example Note]****** Then, the output should be: {"label": [Label], "keywords": [Keywords]}<br><br>Fourth example, if the clinical note is: ******[Example Note]****** Then, the output should be: {"label": [Label], "keywords": [Keywords]}<br><br>Fifth example, if the clinical note is: ******[Example Note]****** Then, the output should be: {"label": [Label], "keywords": [Keywords]}<br><br>Please also find the definition of cognitive decline that may help you to make your analysis: Determining the presence of cognitive decline aimed to identify patients at any stage of cognitive decline, | This template is designed for five-shot prompting. As illustrated in Supplementary **Figure S1**, it includes the task description section, the prompt augmentation section, and the additional task guidance section. |

| | | |
|---|---|---|
| | ranging from SCD to MCI to dementia. Therefore, cognitive decline can be captured by the mention of a cognitive concern, symptoms (e.g., memory loss), diagnosis (e.g., MCI, AD dementia), cognitive assessments (e.g., Mini-Cog) (including patients with normal performance but with a note indicating a cognitive concern), or cognitive-related therapy or treatments (e.g., cognitive-linguistic therapy). We focused on progressive cognitive decline that is likely to be consistent with or lead to MCI. Cases that were less likely progressive (e.g., cognitive function has improved), transient (e.g., temporarily forgetful or occasional memory loss due to medication intake [e.g., codeine]), or reversible (e.g., cognitive function affected soon after some event [e.g., surgery, injury, or stroke]) were considered negative for cognitive decline. We also labeled sections of notes as negative when the record showed broader or uncertain indication of cognitive decline.<br><br>In addition, please also add an explanation for your analysis below the output.<br><br>Now, please analyze this clinical note:******[Note]****** | |
| Template 7 | Using the clinical text provided below, please analyze and determine if there is evidence of cognitive decline in the patient. For your analysis, provide the output in a structured JSON format, which should include the following fields: -'label': Indicate 'Yes' if evidence of cognitive decline is found, or 'No' if not. -'keywords': Provide all distinct words in the clinical text that help you make your judgment.<br><br>Please find the definition of cognitive decline that may help you to make your analysis: Determining the presence of cognitive decline aimed to identify patients at any stage of cognitive decline, ranging from SCD to MCI to dementia. Therefore, cognitive decline can be captured by the mention of a cognitive concern, symptoms (e.g., memory loss), diagnosis (e.g., MCI, AD dementia), cognitive assessments (e.g., Mini-Cog) (including patients with normal performance but with a note indicating a cognitive concern), or cognitive-related therapy or treatments (e.g., cognitive-linguistic therapy). We focused on progressive cognitive decline that is likely to be consistent with or lead to MCI. Cases that were less likely progressive (e.g., cognitive function has improved), transient (e.g., temporarily forgetful, or occasional memory loss due to medication intake [e.g., codeine]), or reversible (e.g., cognitive function affected soon after some event [e.g., surgery, injury, or stroke]) were considered negative for cognitive decline. We also labeled sections of notes as negative when the record showed broader or uncertain indication of cognitive decline.<br><br>Please also keep in mind: 1) short-term symptom like "confusion", "delirium", "change of mental status", "altered mental status" are not considered cognitive decline since they may disappear soon; 2) | This template is an improved version of Template 1, which instructs the LLM to focus on six types of mistakes identified in the 200 training samples. It includes the task description section, an error analysis-based instructions section, and an additional task guidance section, as illustrated in Supplementary **Figure S1**. |

family history of cognitive decline also does not indicate the patient has cognitive decline; 3) swallowing issue is a strong indicator of cognitive decline, especially when combining with other relevant symptoms; 4) depression and anxiety are not considered a sign of cognitive decline; 5) cognitive decline is long-term, and any symptoms that could disappear in a short-term would not be a sign of cognitive decline; 6) cognitive decline related symptoms that are caused by other factors like head injury or bad health condition are not considered a sign of cognitive decline.

In addition, please also add an explanation for your analysis below the output.

Here is the clinical text for analysis: ******[note]******

---

*****The "[note]" represents the actual clinical note section filled in the prompt template; "[Example Note]", "[Label]", and "[Keywords]" represent an example note along with their ground truth label and keywords, which were used for prompt augmentation.

**Table S4.** Preliminary Results of Manual Template Engineering on Ten Training Cases.

| Model | Prompt Template | Percentage of Effective Responses | Execution Time (min) | Estimated Cost | GPT-4 Execution Date |
|---|---|---|---|---|---|
| GPT4-8K ($0.03 per 1000 prompt token. $0.06 per 1000 response token) | Template 1 | 100.0% | 2 | $0.21 | 01/01/2024 |
| Llama2-13B-Q3K-Large ($1.624 per hour) | Template 1 | 20.0% | 12 | $0.32 | - |
| | Template 2 | 80.0% | 12 | $0.32 | - |
| | Template 3 | 30.0% | 8 | $0.22 | - |
| | Template 4 | 50.0% | 12 | $0.32 | - |
| | Template 5 | 40.0% | 14 | $0.38 | - |

**Table S5.** Accuracy of Most Effective Responses from GPT-4 and Llama 2 on Dataset I-S.

| Model | Prompt Template | Percentage of Effective and Parseable Responses | Accuracy | GPT-4 Execution Date |
|---|---|---|---|---|
| GPT4-8K | Template 1 | 99.5% | 86.5% | 12/28/2023 |
| Llama2-13B-Q3K-Large | Template 2 | 86.0% | 52.0% | 12/30/2023 |

**Table S6.** Testing the Impact of Prompt Augmentation on Performance in Dataset I-S.

| Model + Template | Prompt Type | Augmentation Strategy | Accuracy | Precision | Recall | F1 Score | Specificity | Estimated Cost | GPT-4 Execution Date |
|---|---|---|---|---|---|---|---|---|---|
| GPT4-8K + Template 1 | N/A | Baseline (No Augmentation) | 86.5% | 64.7% | 93.6% | 76.5% | 84.3% | $4.49 | 12/28/2023 |
| GPT4-8K + Template 7 | Hard Prompting | Random Selection | 83.0% | 58.44 % | 95.74 % | 72.58% | 79.1% | $11.45 | 12/26/2023 |
| | | Targeted Selection | 84.5% | 61.11 % | 93.62 % | 73.95% | 81.70% | $11.78 | 12/28/2023 |
| | | K-Mean Clustering-Aided Selection | 77.5% | 51.1% | 95.7% | 66.7% | 71.% | $12.56 | 04/14/2024 |
| | Soft Prompting | Dynamic Selection | 85.0% | 62.0% | 93.6% | 74.6% | 82.4% | $12.51 | 01/11/2024 |

**Table S7.** Testing the Impact of Adding Error Analysis-Based Instructions on Performance in Dataset I-S.

| Model | Prompt Template | Accuracy | Precision | Recall | F1 Score | Specificity | Estimated cost | GPT-4 Execution Date |
|---|---|---|---|---|---|---|---|---|
| GPT4-8K | Template 1 | 86.50% | 64.71% | 93.61% | 76.52% | 84.31% | $4.49 | 12/28/2023 |
| | Template 6 | 93.00% | 82.35% | 89.36% | 85.71% | 94.12% | $5.47 | 01/03/2024 |

**Table S8.** Optimized Parameters for Traditional AI Models.

| **Parameter** | **Optimized value** |
|---|---|
| **XGBoost** | |
| TF-IDF normalization | L2 |
| Smooth IDF (+1 to doc frequencies) | True |
| Eta (learning rate) | 0.1 |
| Max depth | 100 |
| Gamma (min split loss) | 0 |
| **Deep Learning** | |
| Word embedding dimensions | 100 |
| Character embedding dimensions | 30 |
| Min word frequency cut-off | 5 |
| LSTM hidden layers | 1 |
| LSTM hidden layer size | 200 |
| CNN hidden layers | 4 |
| CNN hidden layer size | 50 |
| CNN kernel size | 3 |
| CNN padding | 1 |
| Optimizer | SGD |
| Batch size | 10 |
| Learning rate | 0.05 |
| Epochs | 30 |

**Table S9.** Examples of GPT-4's Explanations for Identifying Cognitive Decline from a Long List of Medications.

| Medication Report | GPT-4's Identification | GPT-4 Keywords | GPT4's Explanation |
|---|---|---|---|
| Medication Sig Dispense Refill acetaminophen (TYLENOL) 325 mg tablet Take 3 tablets (975 mg total) by mouth every 8 (eight) hours…... donepezil (ARICEPT) 5 MG tablet Take 1 tablet (5 mg total) by mouth nightly. 90 tablet 3…… sodium fluoride (PREVIDENT) 1.1 % Gel Dose: 1 APPLICATION; Form: Not available; Route: TOP; Frequency: QHS; Directions: 5000 plus; Details: Dispense: 51 Gram(s); Date: 03/07/2016. | The patient has cognitive decline | "donepezil (ARICEPT)" | The patient is taking Donepezil (ARICEPT), which is a medication used to treat symptoms of dementia in people who have Alzheimer's disease. This indicates that the patient may be experiencing cognitive decline. |
| montelukast (SINGULAIR) 10 mg tablet Take 10 mg by mouth nightly. - Oral lidocaine (LIDODERM) 5 % Place 1 patch onto the skin daily…...Inhalation memantine (NAMENDA) 10 MG tablet Take 10 mg by mouth 2 (two) times a day...... - Nasal (C) nitroglycerin (NITROSTAT) 0.4 MG SL tablet [start:9/24/19] Place 1 tablet (0.4 mg total) under the tongue every 5 (five) minutes as needed for chest pain (angina or angina equivalent (nausea)). Resume 9/24/19- pls dispense 4 bottles of 25 mg each - Sublingual (C). | The patient has cognitive decline | "memantine (NAMENDA)" | The patient is taking memantine (NAMENDA), which is a medication used to treat moderate to severe Alzheimer's disease. This suggests that the patient may be experiencing cognitive decline. |

### 3. Supplementary Figures

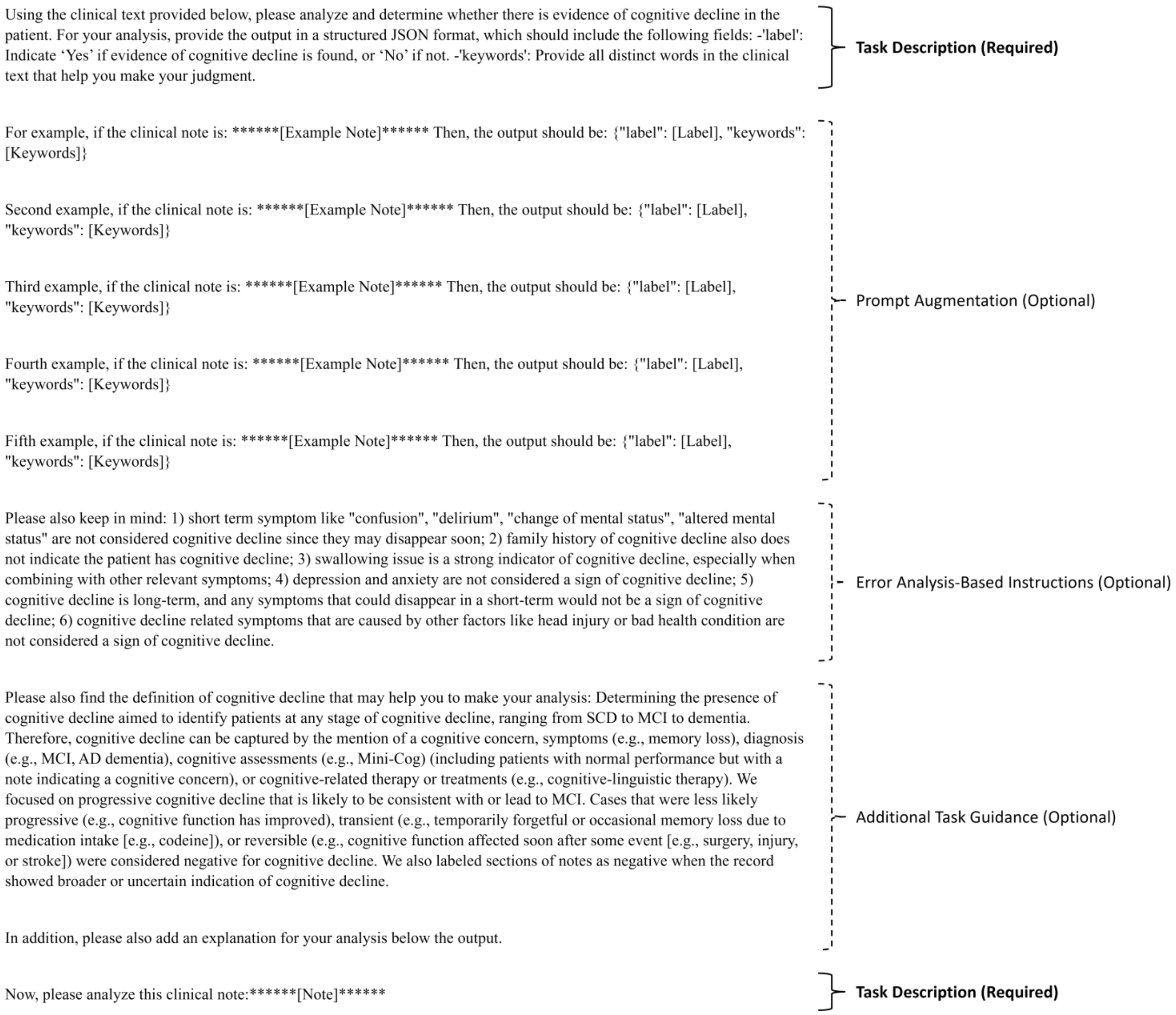


**Figure S1.** Illustration of Prompt Structure.

The structure includes a required section: task description, and optional sections: prompt augmentation, error analysis-based instructions, and additional task guidance. The task description and additional task guidance can be written without using data samples. However, the prompt augmentation and error analysis-based instructions require data samples; prompt augmentation necessitates examples for inclusion in the prompt, and error analysis-based instructions are derived from a summary of incorrectly predicted training samples. Notably, Template 1 in Supplementary **Table S3** includes the task description section and additional task guidance; Templates 2 and 5 include only the task description section; Templates 3 and 4 include both the task description and additional task guidance; Template 6 includes the task description, prompt augmentation, and additional task guidance; Template 7 includes the task description, error analysis-based instructions, and additional task guidance.

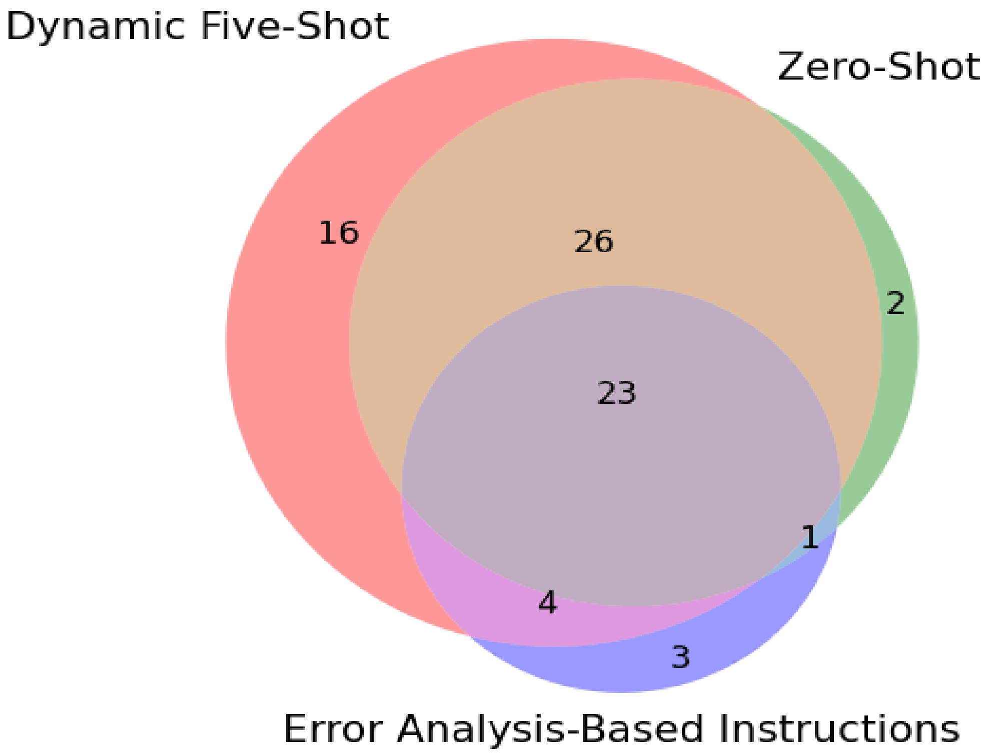


**Figure S2.** Error Profiles for Different Prompting Strategies.

We observed that the error profiles of GPT-4 models with different prompting strategies were less diverse compared to the error profiles among GPT-4, the attention-based DNN, and XGBoost. There were a total of 76 errors. Dynamic five-shot prompting accounted for 69 errors. Notably, 23 (30.3%) of these errors were common across all prompting strategies.
